# Symptom Cluster Profiles among Community-residing Older Adults with Heart Failure: Findings from the U.S. Health and Retirement Study

**DOI:** 10.1101/2024.07.25.24309835

**Authors:** Zequan Wang, Sangchoon JEON, Christine TOCCHI, Samantha CONLEY, Stephen WALSH, Kyounghae KIM, Deborah CHYUN, Nancy Schmieder REDEKER

**Author notes:** Correspondence should be sent to Zequan Wang, PhD, RN University of Connecticut School of Nursing, Storrs, Connecticut 231 Glenbrook Rd, Unit 4026.

## Abstract

**Background:** The incidence of heart failure (HF) rises significantly as people age due to the accumulated influence of risk factors in cardiovascular structure and function. Among older adults with HF, symptoms are manifested in clustered symptoms. Few studies have addressed symptoms specifically in older adults with HF and most have been conducted with small samples. The aims of this study were to (1) describe symptom cluster profiles in older adults with HF in a nationally representative sample of community-dwelling older adults; and (2) determine the associations between demographic and clinical characteristics and membership in symptom cluster profiles.

**Methods:** A secondary analysis was conducted using data from the Health and Retirement Study. Fatigue, shortness of breath, pain, swelling, depressive symptoms, and dizziness were measured. Latent class analysis was used to identify symptom cluster profiles. Bivariate associations and multinomial logistic regression were used to determine the associations between symptom cluster profiles and demographic and clinical characteristics.

**Results:** The sample included 690 participants. Three symptom cluster profiles were identified [high-burden, low-burden, and cardiopulmonary-depressive]. Age, gender, BMI, marital status, alcohol consumption, diabetes, lung disease, and arthritis were significantly different across the three profiles. People in the high-burden and cardiopulmonary-depressive profiles, compared to those in low-burden, had higher odds of having lung disease and arthritis, yet lower odds of higher alcohol consumption.

**Conclusions:** Older adults with HF residing in the community experienced distinct symptom cluster profiles. Research is needed to identify and test targeted interventions for specific symptom cluster profiles.

## BACKGROUND

Heart failure (HF) is a complex clinical syndrome experienced by approximately 26 million adults throughout the world.^1^ The prevalence of HF increases sharply with age due to age-associated changes in cardiovascular structure and function,^2^ and the prevalence of HF among adults aged 60 to 79 years ranges from 4.8% to 6.6%, and it increases to 10.6% to 13.5% for adults aged 80 years and older.^3^ Fatigue, shortness of breath/dyspnea, pain, depressive symptoms, and insomnia symptoms are common among older adults^4–6^ and these symptoms are often associated with advanced age and multimorbidity. These symptoms may also contribute to poor quality of life and function and difficulty with self-care.^7^

Adults with HF demonstrate heterogeneous symptom experiences as evidenced by variations in symptom cluster profiles.^5,8^ While numerous studies addressed isolated symptoms, better understanding of the nature of symptoms and how they co-occur is essential to guide interventions.^8^ Symptom profiles encompass physical and psychological (emotional) domains that may differ depending on the demographic and clinical attributes of the participants.^5,9,10^

Previous studies have indicated that symptoms among adults with HF (age ≥ 18 years) recruited from hospital wards,^11^ outpatient HF clinics,^9,12–16^ and healthcare centers,^17^ occur in clusters or cluster profiles. However, these studies were conducted in specific regions, lacked national representation, and had limited generalizability due to their small sample sizes. Limited studies have been conducted with older adults from a national-wide population, which often provides insights into the comprehensive real-world symptom experience of older adults with HF. Addressing symptom cluster profiles among older adults with a large sample size can help address the unique challenges in managing and preventing HF in an aging population. This includes identifying risk factors and early symptoms/signs of HF, enabling preventive strategies, and ultimately improving the quality of life and adverse outcomes.

The aims of this study are to (1) describe symptom cluster profiles in older adults with HF in a nationally representative sample of community-dwelling older adults in the USA; and (2) determine the associations between demographics, clinical characteristics, and membership in symptom cluster profiles.

## METHODS

### Design

This study was a cross-sectional study of data obtained from the Health and Retirement Study (HRS), conducted by the Institute for Social Research at the University of Michigan. The HRS is a nationally representative longitudinal study of individuals over the age of 50 years,^18^ designed to understand aging at the population level for community-residing, noninstitutionalized adults. Compared to other waves, the HRS 2008 wave included more participants with HF who had subsequent longitudinal data and so, was used as the baseline. This study used de-identified, publicly available data and was deemed exempt from the Institutional Review Board approval of the University of Connecticut.

### Participants and Sample

Participants from the HRS dataset were considered eligible if they resided in the community and were identified as having HF (C048) and comorbid conditions based on the chronic condition codes found in the claims data. We included participants who reported at least one of the following symptoms: (fatigue, pain, shortness of breath, depressive symptoms, and dizziness). We excluded participants who did not report at least one symptom. Participants provided written informed consent prior to the session and verbally agreed to participate in each interview (Health and Retirement Study, 2008c).

### Variables and Measures

The health conditions of participants were evaluated using self-reported health measures in the HRS, and previous studies have confirmed their accuracy and consistency.^18^

Age, gender, race, body mass index (BMI), marital status, educational level, veteran status, smoking status, and alcohol consumption were elicited from the database. The participants’ self-reported age in years was incorporated into the analysis as a continuous variable, while gender was represented by numerical coding: male (1) and female (0). Self-reported weight in pounds and height in inches were converted into kilograms and meters, respectively, to compute BMI in kg/m^2^. BMI was coded and categorized as <18.5 (underweight), 18.5-24.9 (normal weight), 25-29.9 (overweight), 30.0-39.9 (obesity), ≥40.0 (morbid obesity).^19^ Alcohol consumption was defined as “Do you ever drink any alcoholic beverages such as beer, wine, or liquor?”. Participants who answered “yes” were then asked about the number of days per week for alcohol consumption, the number of drinks per day, and how many days they had four or more drinks on occasion (binge drinking). Comorbidities, including hypertension, diabetes, cancer, lung disease, and arthritis, were obtained from the Health Condition questionnaire.

Shortness of breath, persistent swelling in the feet or ankles, fatigue/exhaustion, and dizziness were evaluated during interviews conducted as part of the core surveys.^18^ The response options for each symptom were “yes”, “no”, and “don’t know.” People who answered “yes” were defined by the investigators as experiencing symptoms. Participants were asked about pain, “Are you often troubled with pain?” and participants who answered yes were then asked about pain intensity “How bad is the pain most of the time: mild, moderate, or severe?” Participants who answered yes to the first question and rated the pain as moderate or severe were classified as having significant pain.^20^ These symptoms were selected because they are highly relevant and specific to people with HF.^9,11,21^

HRS investigators assessed depressive symptoms using a short-form eight-item Center for Epidemiological Studies Depression Scale (CES-D). This scale asked participants to indicate the frequency with which they encountered depression-related symptoms in the previous week, with a range of 0-8, and a cutoff point of 4 or more was used to classify depressive symptoms.^22^ The cutoff point for short-form CES-D was determined by HRS, aligning with the established cutoff on the full CES-D.^22^ The short-form CES-D and the original CES-D exhibit comparable symptom dimensions, and the shorter version of CES-D demonstrates strong validity and internal reliability with a Cronbach’s alpha of 0.83.

### Statistical Analysis

Multiple HRS data files were downloaded from the HRS website and cleaned in SAS (v9.4, Cary, NC, 2020). Personal numbers and household numbers were used to identify participants in the 2008 core surveys. Missing values were imputed according to observed items using PROC MI with the EM-algorithm. The study variables were summarized for participants as means ± standard deviations for continuous variables and proportions for categorical variables. The Phi correlation coefficient was used to determine the strength and direction of the association between symptoms.

This study used latent class analysis (LCA), a method that categories individuals into subgroups based on similar symptom experiences among people with HF, utilizing patterns in categorical data.^23^ This person-centered clustering technique helps in identifying distinct groups within the HF population. LCA, as a data-driven approach, does not require any prior hypotheses, and a minimum of 100 participants is needed for a well-identified model.^24^

We performed LCA with the symptoms of fatigue, pain, shortness of breath, swelling, depressive symptoms, and dizziness. The final model was determined based on the relative model fit, including likelihood ratio G^2^, Akaike information criterion (AIC), Bayesian information criteria (BIC), calculated Akaike information criterion (CAIC), and the adjusted BIC. For the fit statistics, the favorable model was based on the lowest AIC and BIC, however, the selection of the final model should depend on fit statistics, clinical interpretability, and the size of each profile. We utilized the PROC LCA (Version 1.3.0) for SAS, developed by the Penn State Methodology Center.^25^ PROC LCA employs a full-information maximum likelihood method to manage missing data and determines symptom cluster profile memberships for participants who lack symptom data.

Chi-square tests and analysis of variance (ANOVA) were conducted to examine the differences in demographic and clinical factors across identified symptom cluster profiles. We examined the residuals for normal distribution in the ANOVA and adjusted for any skewness in the distribution of residuals by applying logarithmic transformation to the variables. Additionally, to understand how demographic and clinical features varied across different symptom cluster profiles, we employed multinomial logistic regression to develop a model. We developed a parsimonious model using stepwise selection of variables that had significant bivariate association with the symptom cluster profiles. Statistical significance was designated as *p* < .05.

## RESULTS

### Demographic and Clinical Characteristics

Table 1 reports the demographic and clinical characteristics of the participants. The sample included 690 individuals [mean age = 74.9 (SD 10.0) years]. Over half of the sample was female (n=390, 56.6%) and a large majority identified as White (n=551, 79.8%). Veterans represented 26.1% of the total sample. The symptom most commonly reported was shortness of breath (n=419, 60.7%), while dizziness was noted as the least frequent symptom (n=213, 30.9%). Table 2 presents the frequency of the symptom and bivariate correlations between the symptoms. All symptoms were correlated with each other (Phi coefficient=0.15-0.34, *p* < .001).

**Table 1.** Demographic characteristics (N=690)

| **Variables** | **Mean (SD)/N (%)** |
| --- | --- |
| **Age** | 74.9 (10.0) |
| **Gender** |  |
| Male | 300 (43.5) |
| Female | 390 (56.5) |
| **Race** |  |
| White/ | 551 (79.8) |
| Black/African American | 113 (16.4) |
| Other/don't know | 26 (3.8) |
| **BMI** |  |
| Under weight (<18.5) | 35 (5.07) |
| Normal weight (18.5-24.9) | 191 (27.7) |
| Overweight (25.0-29.9) | 192 (27.8) |
| Obesity (30.0-39.9) | 209 (30.3) |
| Morbid obesity (>=40.0) | 63 (9.1) |
| **Marital Status** |  |
| Married/partnered | 335 (48.6) |
| Single | 355 (51.4) |
| **Education** |  |
| High school or less | 492 (71.3) |
| College degree/some college | 114 (16.5) |
| Graduate school | 84 (12.2) |
| **Veterans status** |  |
| Yes | 180 (26.1) |
| No | 510 (73.9) |
| **Smoking status** |  |
| Yes | 66 (9.6) |
| No | 624 (90.4) |
| **Alcohol consumption** |  |
| Yes | 179 (25.9) |
| No | 511 (74.1) |
| **Alcohol consumption (number days per week) N=179** | 2.1 (2.5) |
| **Alcohol consumption (number drinks per day) N=103** | 1.7 (0.9) |
| **Alcohol consumption (binge drinking) N=103** | 1.1 (3.2) |
| **Hypertension** |  |
| Yes | 580 (84.1) |
| No | 110 (15.9) |
| **Diabetes** |  |
| Yes | 319 (46.2) |
| No | 371 (53.8) |

| Variables | Mean (SD)/N (%) |
| --- | --- |
| **Cancer** |  |
| Yes | 142 (20.6) |
| No | 548 (79.4) |
| **Lung disease** |  |
| Yes | 232 (33.6) |
| No | 458 (66.4) |
| **Arthritis** |  |
| Yes | 548 (79.4) |
| No | 142 (20.6) |

**Table 2.** Descriptive statistics of symptoms among older adults with heart failure (N=690. )

| Indicator of Latent Status | n, % | Chi-square |  |  |  |  |
| --- | --- | --- | --- | --- | --- | --- |
|  |  | Swelling | Shortness of breath | Fatigue | Depressive symptom | Dizziness |
| **Pain** |  |  |  |  |  |  |
| Yes | 376 (54.5) | **0.24 | **0.20 | **0.30 | **0.15 | **0.23 |
| No | 314 (45.5) |  |  |  |  |  |
| **Persistent swelling** |  |  |  |  |  |  |
| Yes | 381 (55.2) | --- | **0.27 | **0.21 | **0.17 | **0.17 |
| No | 309 (44.8) |  |  |  |  |  |
| **Shortness of breath** |  |  |  |  |  |  |
| Yes | 419 (60.7) | --- | --- | **0.34 | **0.17 | **0.18 |
| No | 271 (39.3) |  |  |  |  |  |
| **Fatigue** |  |  |  |  |  |  |
| Yes | 332 (48.1) | --- | --- | --- | **0.28 | **0.28 |
| No | 358 (51.9) |  |  |  |  |  |
| **Depressive symptoms** |  |  |  |  |  |  |
| Yes | 407 (67.1) | --- | --- | --- | --- | **0.14 |
| No | 200 (32.9) |  |  |  |  |  |
| **Dizziness** |  |  |  |  |  |  |
| Yes | 213 (30.9) | --- | --- | --- | --- | --- |
| No | 477 (69.1) |  |  |  |  |  |

*Note.* \* $p < .05$ ; \*\* $p < .001$ .

### Model Fit Statistics and Membership of Symptom Cluster Profiles

The symptom cluster profiles were best represented by a 3-class model in the LCA because it was parsimonious and presented better profile separation than other classifications. This model exhibited the lowest values in AIC, BIC, cAIC, Adjusted BIC, and it presented a more clinically coherent rationale compared to the 4-class LCA model (Table 3). As reported in Table 4, profile 1 (N=206, 29.9% of the sample) was comprised of “high-burden” symptoms. Participants in the high-burden symptom cluster profile had a high probability of experiencing pain, swelling, shortness of breath, fatigue, depressive symptoms, and dizziness (probabilities ranged from 0.68 to 0.88). Participants in profile 2 (N=117, 17.0% of the sample) had a low probability of experiencing symptoms, and the probability of experiencing symptoms in this “low-burden” profile ranged from 0.00 to 0.38. Participants in profile 3 were comprised of a “cardiopulmonary-depressive” symptom cluster profile. The probability of experiencing symptoms in the cardiopulmonary-depressive symptom cluster profile ranged from 0.00 (pain, dizziness) to 1.00 (shortness of breath).

**Table 3.** Latent analysis fit statistics (N=690)

| Number of<br>statuses | Likelihood<br>Ratio $G^2$ | Degree of<br>Freedom | AIC | BIC | cAIC | Adj.BIC |
| --- | --- | --- | --- | --- | --- | --- |
| 2 | 71.97 | 50 | 97.97 | 156.95 | 169.95 | 115.67 |
| **3** | **52.25** | **43** | **92.25** | **182.99** | **202.99** | **119.48** |
| 4 | 40.2 | 36 | 94.02 | 216.51 | 243.51 | 130.78 |
| 5 | 26.27 | 29 | 94.27 | 248.52 | 282.52 | 140.56 |

**Note.** Boldface values indicate the selected model for this study; AIC=Akaike's information criterion; BIC=Bayesian information criterion. cAIC = consistent Akaike's information criterion; Adj.BIC = Adjusted Bayesian information criterion

**Table 4.** Probabilities of symptoms in each profile-membership from latent class model (N=690)

| Symptoms | Profile 1<br>(High-burden)<br>N=206 (29.86%) | Profile 2<br>(Low-burden)<br>N=117 (16.96%) | Profile 3<br>(Cardiopulmonary-depressive)<br>N=367 (53.19%) |
| --- | --- | --- | --- |
| Pain | **0.85** | 0.07 | 0.00 |
| Swelling | **0.80** | 0.17 | **0.74** |
| Shortness of breath | **0.88** | 0.05 | **1.00** |
| Fatigue | **0.87** | 0.00 | 0.38 |
| Depressive symptoms | **0.87** | 0.38 | **0.81** |
| Dizziness | **0.68** | 0.06 | 0.00 |

**Note.** Boldface values indicate symptoms that characterize cluster profile

Figure 1 illustrates the profiles of symptom cluster profiles for the 3-class LCA model. The y-axis represents the probability of symptoms, while the x-axis shows indicator variables used for the LCA model. The three lines represent symptom patterns for the three symptom cluster profiles. Crossing was observed among the three lines, suggesting that the exhibited symptoms in older adults with HF are likely associated with more than one cluster profile.

**Figure 1.**
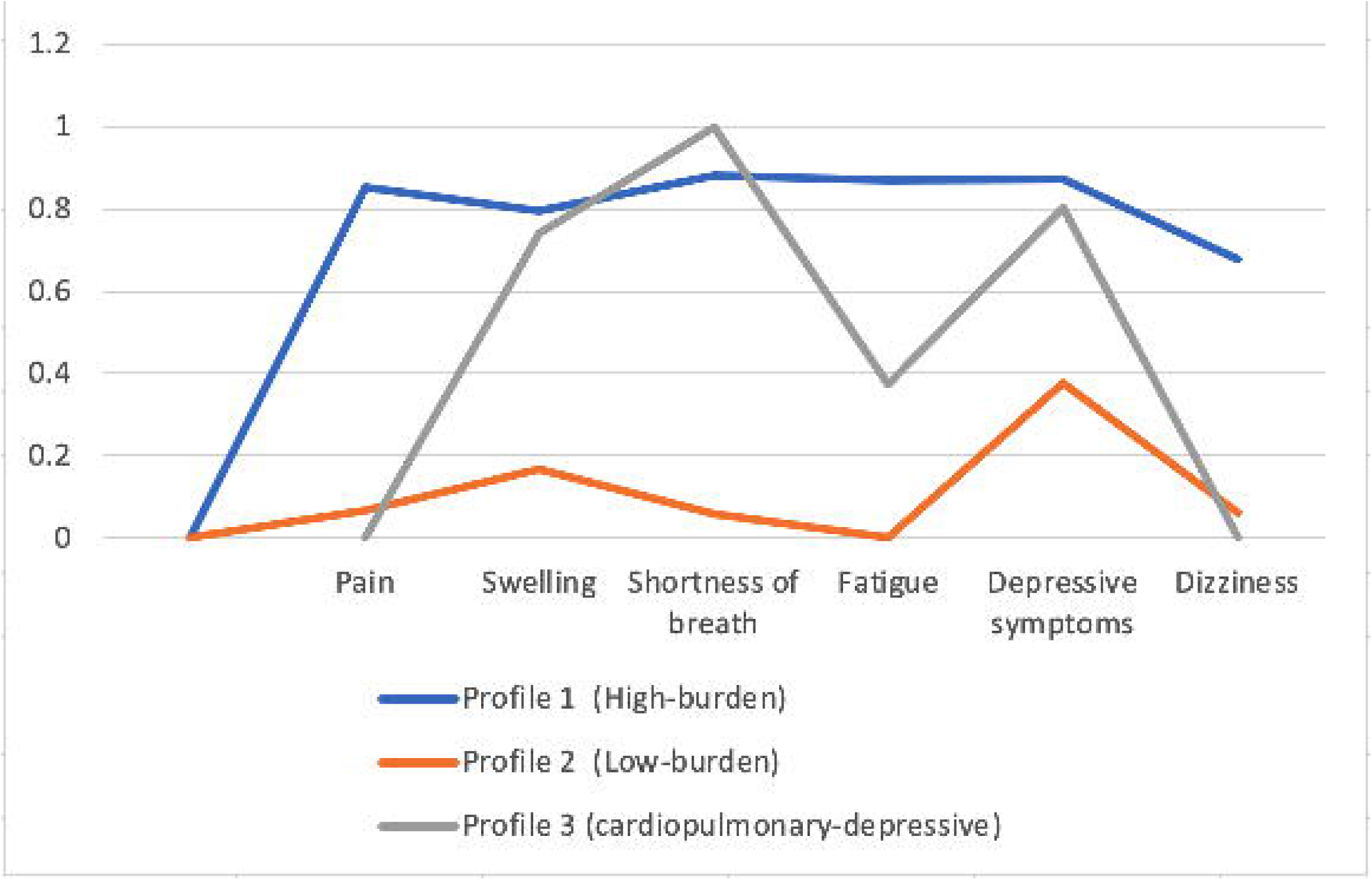
Symptom cluster profiles of latent classes

### Bivariate Analysis and Multinomial Logistic Regression of Symptom Cluster Profiles

We also examined the differences in participants’ demographic and clinical characteristics across the three symptom cluster profiles (Table 5). Age, gender, BMI, marital status, veteran status, alcohol consumption, diabetes, lung disease, and arthritis had statistically significant differences across three symptom cluster profiles (*p* < .05). Participants in profile 3 (cardiopulmonary-depressive) were the oldest (76.4 years ± 10.0) and were more likely to be married or partnered (48.0%), to be veterans (27.5%), and to be male (46.3%), compared to those in profile 1 (high-burden) and 2 (low-burden). Most participants in the high-burden profile were overweight (25.7%), obese (36.4%), or morbidly obese (14.6%). Over one-quarter of participants in the cardiopulmonary-depressive profile were overweight (25.9%) or obese (28.3%).

**Table 5.** Comparison of the three symptom cluster profiles on demographic and clinical (N=690)

| **Variables** | **Profile 1**<br>**All-high**<br>**(N=206)** | **Profile 2**<br>**All-low**<br>**(N=117)** | **Profile 3**<br>**Cardiopulmonary-**<br>**depressive**<br>**(N=367)** | **Differences**<br>**P-value** |
| --- | --- | --- | --- | --- |
| **Age (years)** | 72.2±9.9 | 75.1±9.7 | 76.4±10.0 | **<.001 |
| **Gender** |  |  |  | *0.01 |
| Male | 71 (34.5%) | 59 (50.4%) | 170 (46.3%) |  |
| Female | 135 (65.5%) | 58 (49.6%) | 197 (53.7%) |  |
| **Race** |  |  |  | 0.37 |
| White/Caucasian | 159 (77.1%) | 95 (81.2%) | 297 (80.9%) |  |
| Black/African American | 35 (16.9%) | 20 (17.1%) | 58 (15.8%) |  |
| Other/don't know | 12 (5.8%) | 2 (1.7%) | 12 (3.3%) |  |
| **BMI** |  |  |  | **<.001 |
| Under weight (<18.5) | 8 (3.9%) | 5 (4.3%) | 22 (5.9%) |  |
| Normal weight (18.5-24.9) | 40 (19.4%) | 30 (25.5%) | 121 (32.9%) |  |
| Overweight (25.0-29.9) | 53 (25.7%) | 44 (37.6%) | 95 (25.9%) |  |
| Obesity (30.0-39.9) | 75 (36.4%) | 30 (25.6%) | 104 (28.3%) |  |
| Morbid obesity (≥40.0) | 30 (14.6%) | 8 (6.8%) | 25 (6.8%) |  |
| **Marital status** |  |  |  | *0.03 |
| Married and partner | 90 (43.7%) | 69 (58.9%) | 176 (47.9%) |  |
| Others | 116 (56.3%) | 48 (41.0%) | 191 (52.0%) |  |
| **Education** |  |  |  | 0.05 |
| High school or less | 160 (77.7%) | 78 (66.7%) | 254 (69.3%) |  |
| College and above | 46 (22.3%) | 39 (33.3%) | 113 (30.7%) |  |
| **Veteran Status** |  |  |  | *0.03 |
| Yes | 41 (19.9%) | 38 (32.5%) | 101 (27.5%) |  |
| No | 165 (80.1%) | 79 (67.5%) | 266 (72.5%) |  |
| **Smoking status** |  |  |  | 0.93 |
| Yes | 21 (10.1%) | 11 (9.4%) | 34 (9.3%) |  |
| No | 185 (89.8%) | 106 (90.6%) | 333 (90.7%) |  |
| **Alcohol Consumption** |  |  |  | **<.001 |
| Yes | 38 (18.5%) | 52 (44.4%) | 89 (24.3%) |  |
| No | 168 (81.6%) | 65 (55.6%) | 278 (75.8%) |  |
| **Alcohol consumption**<br>**(number days per week)** | 1.7±2.4 | 1.9±2.3 | 2.3±2.7 | 0.35 |
| **Alcohol consumption**<br>**(number drinks per day)** | 1.9±1.2 | 1.5±0.8 | 1.8±0.9 | 0.24 |
| **Alcohol consumption**<br>**(binge drinking)** | 1.9±5.8 | 0.5±1.4 | 1.1±2.8 | 0.29 |
| **Hypertension** |  |  |  | 0.08 |
| Yes | 183 (88.8%) | 96 (82.1%) | 301 (82.0%) |  |
| No | 23 (11.2%) | 21 (178.0%) | 66 (18.0%) |  |
| **Diabetes** |  |  |  | *0.01 |
| Yes | 115 (55.8%) | 49 (41.9%) | 155 (42.2%) |  |
| No | 91 (44.2%) | 68 (58.1%) | 212 (57.8%) |  |
| **Cancer** |  |  |  | 0.11 |
| Yes | 52 (25.2%) | 19 (16.2%) | 71 (19.4%) |  |
| No | 154 (74.8%) | 98 (83.8%) | 296 (80.6%) |  |
| **Lung disease** |  |  |  | **<.001 |
| Yes | 110 (53.4%) | 18 (15.4%) | 104 (28.3%) |  |
| No | 96 (46.6%) | 99 (84.6%) | 263 (71.7%) |  |
| **Arthritis** |  |  |  | **<.001 |
| Yes | 183 (88.8%) | 79 (67.5%) | 286 (77.9%) |  |
| No | 23 (11.2%) | 38 (32.5%) | 81 (22.1%) |  |

*Note:* \* $p < .05$ ; \*\* $p < .001$ .

Almost one-quarter of participants in the cardiopulmonary-depressive profile had a history of alcohol consumption (24.3%). The average days/week of alcohol consumption per week (2.3 days ± 2.7) was higher compared to profiles 1 and 2. Participants in the cardiopulmonary-depressive profile were more likely to report hypertension (82.0%), diabetes (55.8%), and arthritis (77.9%). However, having lung disease was the most common in participants in the high-burden symptom cluster profile (53.4%).

Table 6 presents the results of the multinomial logistic regression. People in profile 1 (high-burden) and profile 3 (cardiopulmonary-depressive), compared to those in profile 2 (low-burden), had higher odds of having lung disease and arthritis (*p* < .001). People in the high-burden and cardiopulmonary-depressive symptom cluster profiles were less likely to consume more alcohol compared to those in the low-burden profile (*p* < .001).

**Table 6.**
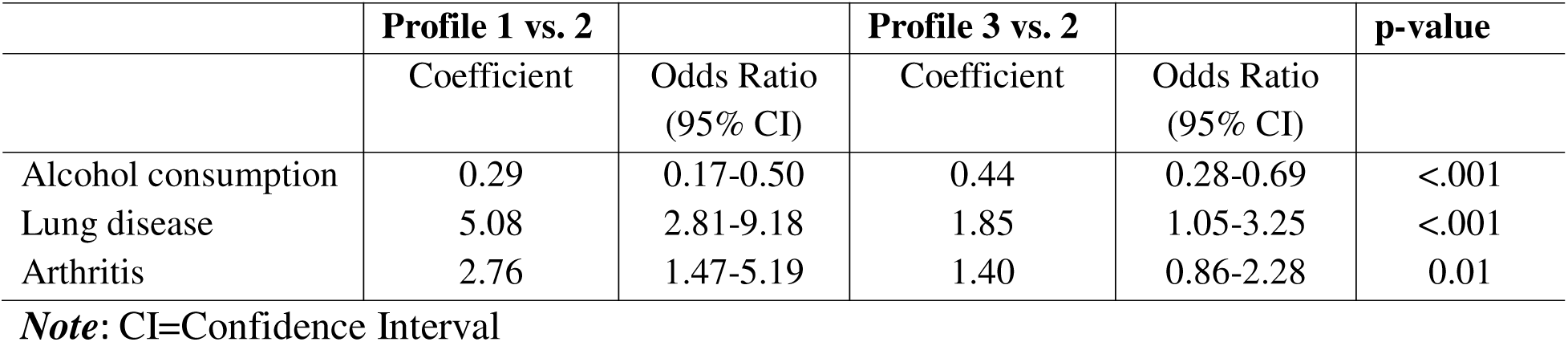
Multinomial logistic model.

|  | **Profile 1 vs. 2** |  | **Profile 3 vs. 2** |  | **p-value** |
| --- | --- | --- | --- | --- | --- |
|  | Coefficient | Odds Ratio<br>(95% CI) | Coefficient | Odds Ratio<br>(95% CI) |  |
| Alcohol consumption | 0.29 | 0.17-0.50 | 0.44 | 0.28-0.69 | <.001 |
| Lung disease | 5.08 | 2.81-9.18 | 1.85 | 1.05-3.25 | <.001 |
| Arthritis | 2.76 | 1.47-5.19 | 1.40 | 0.86-2.28 | 0.01 |

*Note:* CI=Confidence Interval

## DISCUSSION

To our knowledge, this is the first study that evaluated associations between demographic and clinical factors and symptom cluster profiles in a nationally representative sample of community-residing older adults with HF. It extends previous symptom cluster studies that included more geographically localized samples recruited from health care centers and had smaller samples. Two of the identified symptom cluster profiles (high-burden and low-burden), are characterized by either consistently high or low physical and psychological symptoms and are consistent with the symptom cluster profiles identified in previous studies.^5,9,26^ However, the membership in the high-burden and low-burden profiles varies depending on which symptoms were selected. In our study, most participants (53.2%) belonged to the cardiopulmonary-depressive symptom cluster profile, which is similar to those reported in the literature that found integrated physical and psychological symptoms in patients with HF.^11,17,27^

Compared with the low-burden symptom cluster profile, it is not surprising that comorbidity was associated with membership in the high-burden and cardiopulmonary-depressive symptom cluster profiles. Notably, we found that the risk of lung disease for high-burden symptom cluster profiles is about 5 times higher than that of low-burden symptom cluster profiles. Therefore, assessing CRP and fibrinogen would add additional insight into the risk of lung disease among those with severe HF. Elevated inflammatory markers, such as C-reactive protein (CRP) and fibrinogen, perhaps are the most prominently cited risk factors to explain the associations between severe HF and pulmonary dysfunction.^28^ In addition to lung disease and arthritis, previous studies have also found an association between diabetes and emotional symptom cluster (depressed, worry, difficult concentration) in patients with HF.^29^ Hypertension, atrial fibrillation, and diabetes were significant predictors of being in the high distress profile (severe physical and psychological symptoms) compared to the low distress profile.^9^ HF, especially HF preserved ejection fraction (HFpEF), is associated with a high prevalence of comorbidities.^30^ Conditions, such as hypertension (65%), atrial fibrillation (45%), diabetes (40%), and chronic obstructive pulmonary disease (40%), are commonly observed in older adults with HF. These comorbidities, in turn, exacerbate the progression and clinical presentation of HF, forming a vicious cycle. The reciprocal relationships can be particularly challenging among older adults and are considered a true geriatric syndrome.^30^ Age-related changes and cumulative comorbidities can be a challenge for older adults to distinguish whether symptoms are arising from HF itself or from its comorbidities.^31^

The prevalence of depressive symptoms was quite pronounced. Depressive symptoms were also manifest in two distinct symptom cluster profiles. This may be because veterans represented over a quarter of the total participants, and over half of these veterans were categorized within the cardiovascular-depressive symptom cluster profile. Deployed military personnel were twice as likely to suffer depression than non-deployed military personnel.^32^ However, previous HRS studies have shown that veterans and non-veterans had a comparable prevalence of depression and anxiety symptoms, and these emotional symptoms vary among veteran cohorts.^33^ Veterans have reported a reluctance to seek care for fear of stigma and negative consequences, which could result in fear of admissions of depressive symptoms among this population.^34^ Given the high prevalence and intricate nature of depressive symptoms among community-residing older adults with HF, particularly veterans, it is crucial for clinicians and health care providers to prioritize the assessment and management of depression and its symptoms.

Older adults with HF who consume alcohol were less likely to belong to high-burden and cardiopulmonary-depressive symptom cluster profiles. One reason might be that older people with high-burden or cardiopulmonary-related symptoms are more aware of the severity of their condition and the impact of alcohol on their daily lives and were following medical advice to avoid alcohol consumption This may also be attributed to several considerations, such as physiological effects, lifestyle modifications, symptom burden awareness, and patient education.^35^ Consequently, they may be more motivated to adopt lifestyle modifications and prioritize medication/lifestyle adherence, such as limiting alcohol intake, to alleviate symptoms.^35,36^ Additionally, current evidence shows that low amounts of alcohol consumption are safe and beneficial for the cardiovascular system.^36^ However, alcohol consumption can lead to either an elevated or reduced cardiovascular risk, depending on the amount consumed, frequency of drinking, and the pattern of consumption, whether it involves heavy/binge drinking or irregular intake.^37^

It is important to note that these findings are nuanced and not uniform across all symptom profiles. Though our limited sample size prevented us from finding statistically significant results regarding drinking frequency and consumption patterns across different symptom cluster profiles, we did observe that, among older adults with HF who consumed alcohol, those in the cardiopulmonary-depressive profile consumed alcohol more frequently per week. Meanwhile, individuals in the high-burden profile showed a higher tendency for binge drinking compared to other groups. Even though the percentage of individuals with alcohol consumption in the high-burden profile is relatively lower, their average number of drinks per day and frequency of binge drink is much higher, indicating they tend to engage in heavier alcohol use compared to other profiles. The possible reason that alcohol might exacerbate high-burden or cardiopulmonary-depressive symptoms may stem from a combination of direct and indirect physiological effects. These effects increase the risk of dilated cardiomyopathy, elevated blood pressures, interference with medications, promoting fluid retention, triggering cardiac arrhythmias and mitochondrial dysfunction in circulation.^38^

Both physical and psychological/emotional aspects of HF can be addressed through a combination of medication, psychotherapy, and lifestyle changes.^39^ This may help healthcare providers identify the specific symptom cluster profiles that are most affected by depressive symptoms and tailor the treatment accordingly. Several interventions that combine symptom management support and self-care management can assist people with HF in recognizing, responding to, and managing multiple symptoms.^40^ Home-based inspiratory muscle training (IMT) has been found to be a safe, flexible, and adjuvant treatment of pharmacological interventions in reducing fatigue and dyspnea in patients with HF.^41^ Motivational interviewing (face-to-face sessions plus telephone calls) is another effective strategy to reduce discomfort, dyspnea, and early and subtle edema in patients with HF.^42^ Few interventions have intentionally targeted symptom clusters/profiles in people with HF. Future studies should consider developing and examining the effectiveness of interventions aimed at reducing the incidence of specific symptom clusters/profiles for older adults with HF. Cognitive training combined with exercise, self-care, and electrical muscle stimulation may be effective as a non-pharmacological intervention for older adults with HF.^43^

The strength of this study is its large sample size which helps gain insights of symptom cluster profiles among community-residing older adults. Instead of directly assigning people to specific profiles, this study used probabilistic class assignments, allowing for a better evaluation of differences between profiles and the impact of predictive factors on profile assignments. Nevertheless, this study had several potential limitations. Firstly, its cross-sectional nature precludes drawing conclusions on causality and weakens the dynamic analysis of how demographic and clinical factors influence symptom cluster profiles in older adults. Secondly, some symptoms, such as shortness of breath and fatigue, were identified using one single question from self-reported surveys, which may affect the accuracy and validity of the results. Due to limited data in the HRS, sleep or insomnia symptoms were not included. However, most people with HF report poor sleep, and 50% report insomnia symptoms, including difficulty initiating sleep, maintaining sleep, or awakening too early in the morning.^44^ Lastly, the limited diversity in race and ethnicity, combined with limited clinical data (e.g., New York Heart Class), might impact the accurate characterization of symptoms.

HF clinicians should carefully evaluate older adults with HF who reside in communities for the presence of multiple co-occurring symptoms. It is especially important to assess and manage depressive symptoms among veterans as these symptoms may present within high-burden and cardiovascular-depressive symptom cluster profiles. Clinicians and health care providers should also educate older adults with HF, particularly those experiencing low-burden symptoms, about the risks associated with alcohol consumption and the potential negative consequences. Symptoms may change and deterioration can occur in later life, and various risk factors may be associated with dynamic symptom trajectory among this vulnerable population. Future research is needed to determine the longitudinal trajectory of symptom cluster profiles and examine the extent to which risk factors can predict changes in symptom cluster membership.

## CONCLUSIONS

In conclusion, among community-residing older adults with HF, we identified three distinct symptom cluster profiles: high-burden, low-burden, and integrative physical and emotional symptom cluster profiles. Lung disease and arthritis are associated with the high-burden symptom cluster profiles, while alcohol consumption is associated with the low-burden symptom cluster profiles. There is a need to examine these symptom cluster profiles longitudinally and pinpoint older adults at risk for adverse outcomes. Targeted symptom cluster profile intervention should also be explored.

## List of abbreviations

HF: Heart failure
HRS: Health and retirement study
BMI: Body mass index
CES-D: Center for Epidemiological Studies Depression Scale
LCA: latent class analysis
AIC: Akaike information criterion
BIC: Bayesian information criteria
CAIC: calculated Akaike information criterion
ANOVA: Analysis of variance
HFpEF: HF preserved ejection fraction

## Acknowledgements

Not applicable

## Author contributions

Z.W. designed the study and drafted the manuscript. Z.W., S.J., N.S.R., and S.W. contributed to the methodology. Z.W. performed statistical analyses. All authors contributed to the interpretation of the results and read and approved the final manuscript.

## Data Availability

The datasets generated and/or analyzed in this current sutdy are available from the U.S Health and Retirement Study (https://hrs.isr.umich.edu/about)

## Declarations

### Ethics approval and consent to participate

This study used de-identified, publicly available data and was deemed exempt from the Institutional Review Board approval of the University of Connecticut. Participants provided written informed consent prior to the session and verbally agreed to participate in each interview (Health and Retirement Study, 2008c).

### Consent for publication

No applicable.

